# Association of Outpatient Crisis Services with Emergency Department Revisits in Suicidal Youth

**DOI:** 10.64898/2026.11.09.26367462

**Authors:** William E. Copeland, Janet R. Cummings, Phoebe K. McCutchan, Nikolay Braykov, Naveen Muthu, Xiaotao Jing, Afrin Jahan, Cristian Ramos, Laura J. Dilly, Kathleen H. Krause, Belise Livingston-Burns, Lisa R. Schneider, Marcus Graham, Gargi Mukherjee, Hillary M. Henderson, James J. Hudziak, John Constantino

## Abstract

**Background and Objective:** Pediatric emergency department (ED) visits for suicidal ideation and behavior have increased substantially, highlighting the need for scalable outpatient interventions that reduce recurrent ED use. We tested whether referral to outpatient crisis services was associated with fewer mental health-related ED revisits, hypothesizing that referred youth would have lower revisit rates than those receiving usual community care.

**Methods:** We conducted a quasi-experimental study of 3,125 youth presenting with suicidal ideation or behavior to a large academic children’s health system between October 1, 2023, and August 31, 2025. Youth were referred to outpatient crisis services or community treatment (usual care) based on slot availability and clinician discretion. Propensity score modeling balanced demographic, clinical, and prior treatment characteristics between groups.

**Results:** Among 3,125 qualifying index ED visits, 763 (24.4%) youth were referred to crisis services and 461 (14.8%) received ≥1 crisis visit within 30 days. In intent-to-treat analyses, referral was associated with fewer mental health-related ED revisits at 90 days (adjusted incidence rate ratio [aIRR], 0.67; 95% CI, 0.56–0.81) and 180 days (aIRR, 0.68; 95% CI, 0.59– 0.80). Associations were strongest among youth requiring short-term hospitalization (aIRR, 0.46; 95% CI, 0.36–0.58) and those with moderate-to-high suicide risk (aIRR, 0.49; 95% CI, 0.39– 0.61). Findings were consistent in as-treated analyses.

**Conclusion:** Outpatient crisis services were associated with substantially fewer mental health-related ED revisits among suicidal youth, with the largest reductions among youth with greater clinical acuity.

## Introduction

Youth visits to the emergency department for mental health concerns are rising. In 2011, 7.7% of pediatric ED visits were mental health-related, but this nearly doubled to 13.1% by 2020 with an average annual percent change of 8%.^1^ Follow-up with mental health treatment after discharge remains concerningly low, with only 31.2% of children having an outpatient visit within 7 days and only 55.8% within 30 days.^2^ Consequently, 26.5% of youth with a mental health-related ED visit returned for an acute care encounter within 6 months.^2^ The ED increasingly serves as a safety net for youth with mental health needs but this comes at a significant financial cost to health systems and often fails to provide youth in crisis with optimal longer-term outpatient care.^3^

The greatest increase in mental health-related ED visits from 2011 to 2020 was for suicide-related visits, which rose from 0.9% to 4.2% of all pediatric ED visits.^1^ This increase was driven, in part, by ED visits for suicide attempt and intentional self-harm in adolescent girls, which were double that of any other age/gender group.^4^ Of youth with a suicide-related ED visit, 11.7% returned for an acute care encounter within 7 days and 25.7% returned within 6 months.^2^ Revisits were associated with indicators of higher burden of need—public insurance, multiple psychiatric comorbidities; indicators of advantage, including a higher Child Opportunity Index score; and geographic location, such as residing in an urban location and in close proximity to an ED.^5^

How can a health system reverse these trends and reduce costly ED revisits in youth with suicidal thoughts and behaviors? Access to outpatient services in a timely manner can reduce revisits short-term within the first week,^2^ but beyond that, the results are equivocal.^2,6,7^ Proposed explanations for these mixed findings include varying availability and quality of follow-up care, unaccounted for psychosocial complexity, differences in disease severity, or inconsistent definitions of diagnostic groupings. However, it may also be that the standard outpatient care – beneficial though it may be – is simply unable to prevent the type of crises that result in ED revisits.^2,6,8^

Further research must test whether high-quality, evidence-based outpatient care can reduce ED recidivism for children with suicidality. In 2023, Children’s Healthcare of Atlanta initiated expenditure of an institutional resource to subsidize the cost of high-quality outpatient behavioral health services to a large cohort of population-representative youth within its system. As an early priority, a crisis recovery services program was established to implement rapid, empirically supported outpatient care for youth with suicidality. Need for these services exceeded the capacity of the new resource, resulting in some youth being referred internally to receive the new crisis services (treatment) and others to care as usual in the community. This program evaluation examines whether children referred to crisis services after presenting to the ED for suicidality were less likely to revisit the ED as compared to their peers receiving care as usual.

## Methods

Children’s Healthcare of Atlanta is a national leader in pediatric emergency department utilization, operating three children’s hospitals and managing more than 230,000 ED visits annually. The three hospitals receive 25% of all emergency department encounters of children in behavioral crisis in the State of Georgia. For this program evaluation, emergency department visit data were combined with electronic health record data on outpatient mental health visits to the Behavioral Mental Health (BMH) outpatient clinics. The Children’s Healthcare of Atlanta IRB determined this retrospective review of clinical data as non-research, conducted for the purpose of quality improvement and program evaluation.

### Study Population

Between October 1^st^ 2023 to August 31, 2025, over 5,500 emergency department (ED) visits were designated as mental health-related and included emergency assessments by LCSW mental health specialists (MHS) as well as emergency pediatricians. In 3,474 of these visits, suicidal thoughts and behaviors were identified as either 1) the primary or secondary referral reason or 2) the primary or secondary clinical impression reported by the MHS in the ED. To focus on the novel impact of crisis services received after the index visit, we only included ED visits in which the child had not had a mental health-related ED visit in the previous 180 days. Also, visits in which suicide risk was initially suspected but later judged not to be present were excluded (n=26). These criteria resulted in 3,125 total qualifying index visits.

At the index ED visit, patients with high symptom severity and/or significant safety concerns were commonly transferred to an inpatient psychiatric setting for stabilization prior to engaging in either treatment condition using a legal process known in Georgia as a 1013 order. For non-hospitalized patients, the emergency physician could either refer the child internally to the system’s new outpatient crisis recovery program (i.e., treatment group) or to treatment in the community (i.e., Care as Usual). Internal referral was based on slot availability and emergency clinicians’ discretion.

### Exposure

Outpatient crisis services at Children’s consists of two primary options intended to provide rapid access (ideally within 72 hours) to high-quality, short-term recovery: (1) empirically-based psychopharmacology services; and (2) intensive outpatient psychotherapy using the Collaborative Assessment and Management of Suicidality (CAMS) framework,^9^ supplemented by weekly dialectical behavior therapy skills group therapy. CAMS is an intensive short-term evidence-based intervention designed to engage individuals in a collaborative therapeutic process that directly targets ‘drivers’ of suicidal thinking and behavior. Prior studies in adults have indicated that CAMS effectively reduces suicidal ideation and overall distress, while promoting a sense of hope and retention in care;^10^ a pilot study of the adapted version for youths (CAMS-4Teens) has also yielded promising findings.

### Outcome Measures

Two count variables were created for the total number of ED revisits for a mental health concern in each group within 90 or 180 days of the index ED visit. Three to six months were chosen as targets as revisits within this period are more likely to be clinically associated with the treatment at the index visit. We excluded revisits occurring within 72 hours of the index visit as they were considered part of the same clinical episode.

### Covariates

Sociodemographic and clinical variables were extracted from the electronic medical record to adjust for potential group differences. Demographic variables included sex at birth, age, race/ethnicity (i.e., Black, Hispanic/Latino, White, Other), insurance status (i.e., Medicaid, Private, Self-pay, and Other), distance from geocoded home address to Children’s hospitals (i.e., more or less than 50 miles from the hospital), preferred language (i.e., English, Spanish, or Other), and the Child Opportunity Index of geocoded home address. Clinical variables included whether or not the ED visit resulted in transfer to inpatient-psychiatric treatment, the Children’s campus where the index ED visit occurred, suicidality acuity based upon results from the Ask Suicide-screening Questions (ASQ) tool^14^ and the mental health specialists’ interview (three levels: red equals highest risk, typically current active suicidal ideation, yellow equals moderate risk, and green equals low suicide risk), family report of maltreatment, history of child welfare involvement with the family (yes/no), and quarter of the year the visit occurred. Additional dichotomous variables indicated prior mental health ED visits to Children’s hospitals (before the last 180 days), any prior mental health inpatient visits to Children’s hospitals, and any prior mental health outpatient visits within the Children’s health system. Parents also reported on mental health services the child received outside of the Children’s system including outpatient (yes/no), inpatient (yes/no), and medication related (yes/no).

### Analytic Plan

Propensity score modeling was used to balance variables related to sociodemographic and clinical characteristics between groups.^15–17^ This balance ensures that potential differences in treatment and care as usual referrals based on slot availability and clinical discretion are controlled. Covariate balance was tested by comparing the standardized mean differences between groups on all covariates after applying the inverse probability of treatment weights. The goal was to reduce standardized mean differences to less than 0.10 between the groups. Any covariates without adequate balance were also included as covariates in the final model estimating the treatment effect. Weighted negative binomial regressions were used to obtain an estimate of the association between referral to crisis services and the total count of ED revisits in each group at 90 or 180 days.

All analyses were performed in R 4.4.0 with adapted code from package “WeightIt” for all outcome regression models.^18^ Findings were considered statistically significant using a two-tailed test at *P* < 0.05, with incident rate ratios and confidence intervals presented for negative binomial regression models.

## Results

Between October 1^st^ 2023 to August 31^st^ 2025, 3,125 ED visits met our criteria as a qualifying index ED visit for suicidal thoughts and behaviors. Of these, 11% had a return visit to the ED within 90 days and 15% had a return visit within 180 days. This compares to return rates of 12% and 16% in the preceding year before study began. Of the 3,125 index visits, 763 youth (24%) were referred to crisis services (i.e., treatment group). 64% received a CAMS referral and 55% received a psychopharmacology referral, with 19% of youth receiving referrals to both services. Of these, 461 completed at least 1 visit within 30 days following their index ED visit and 298 completed at least 2 visits within 30 days. In the treatment group, the mean number of revisits within 90 days was 0.10 (SD=0.35) and within 180 days was 0.14 (SD=0.21). In care as usual, the mean number of revisits in 90 days was 0.17 (SD=0.62) in 90 days and 0.27 (SD=0.37) in 180 days. **Table 1** displays descriptive information for both the treatment and care as usual groups. Both groups were majority female (62-65%), majority Black (51-58%), and the majority were insured with Medicaid (51-59%).

**Table 1.**
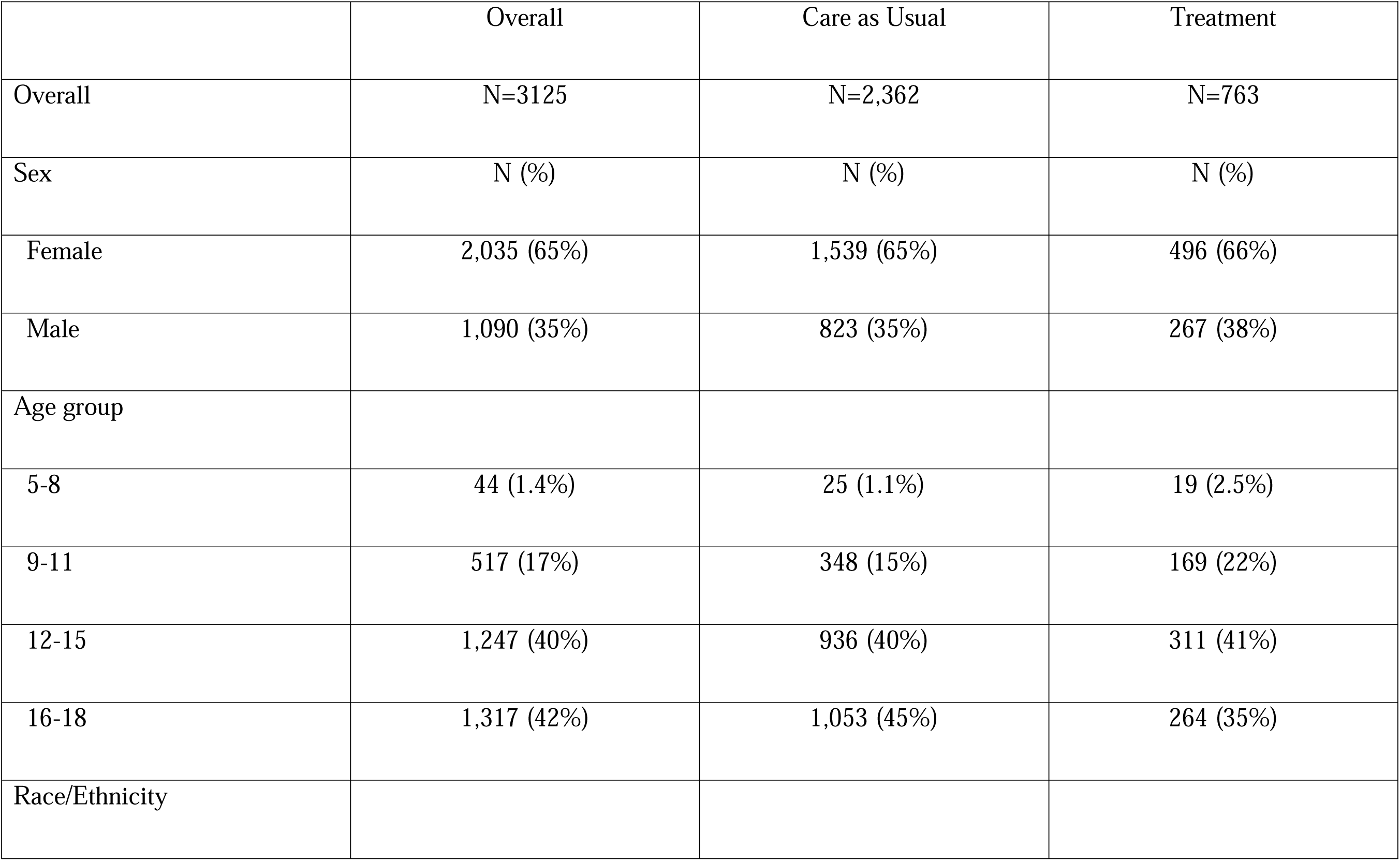
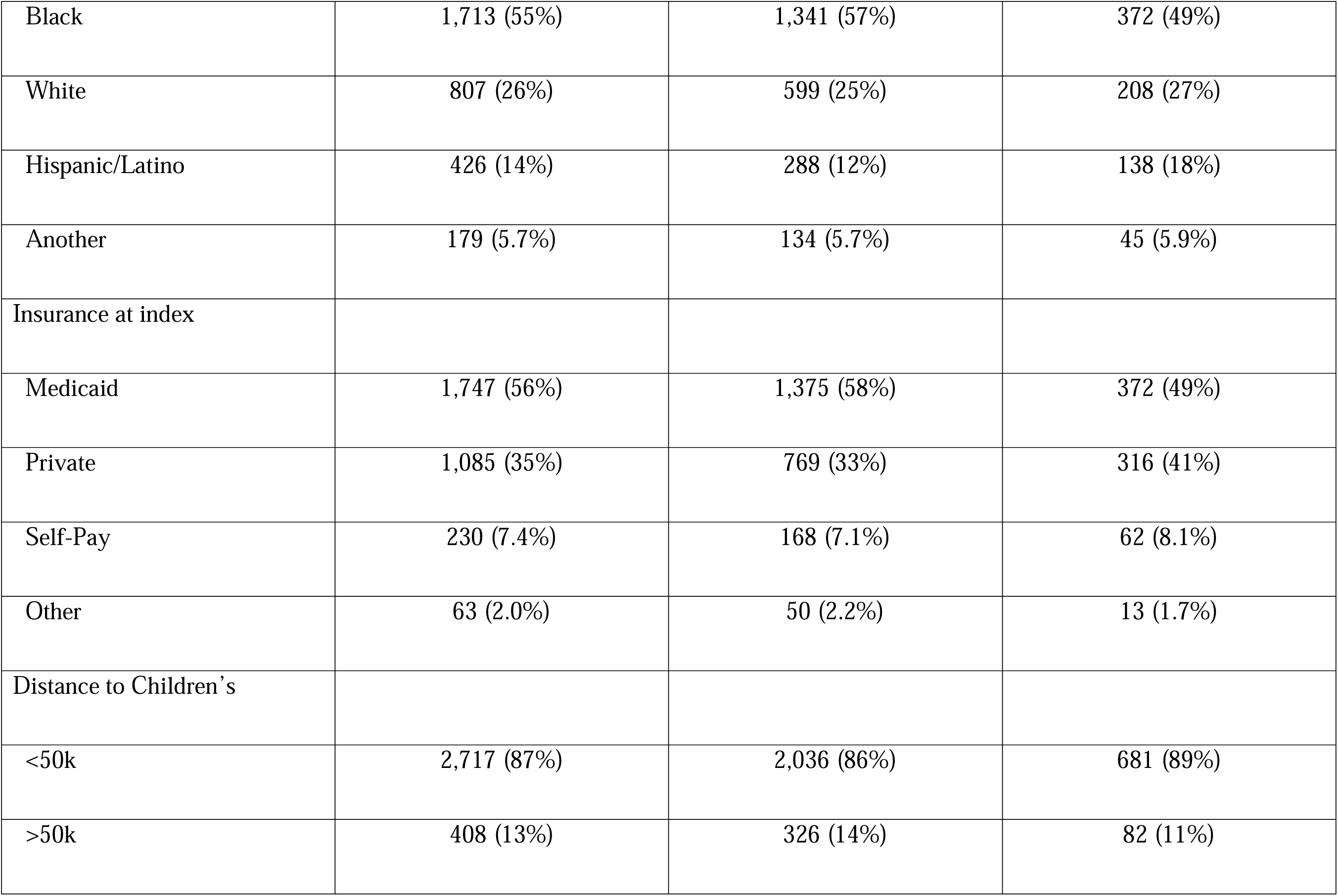
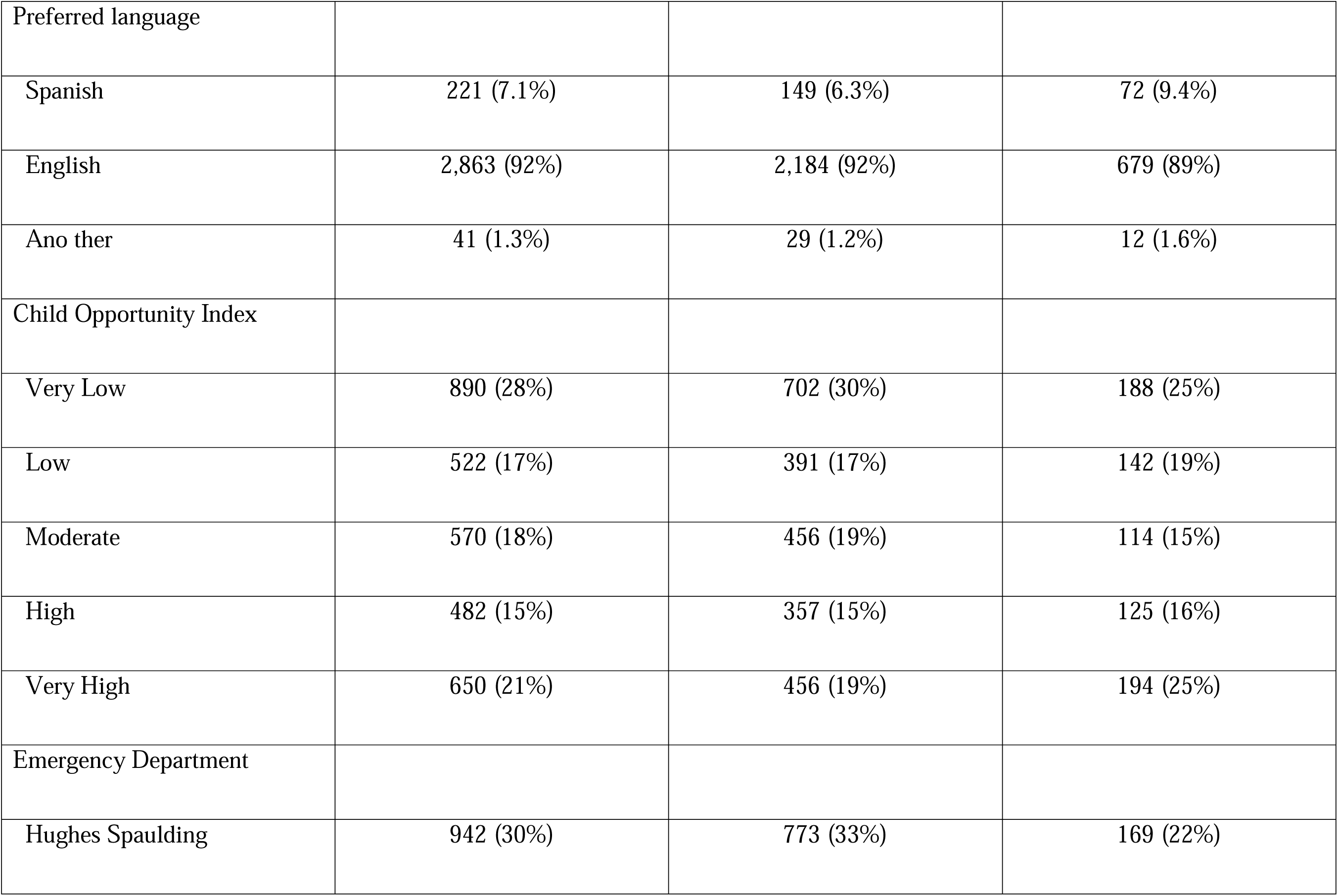
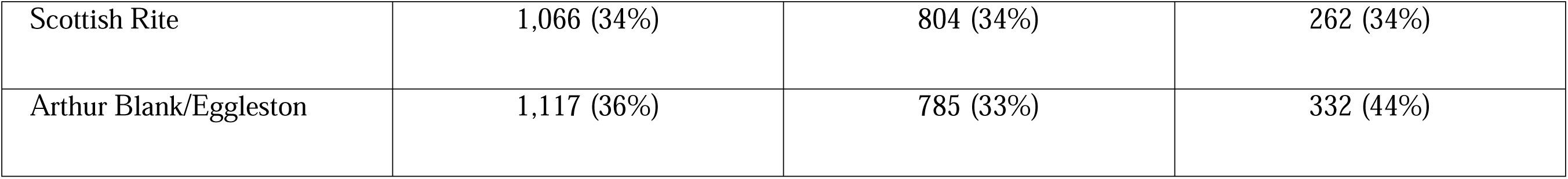
Descriptive statistics for Children’s crisis and care as usual groups.

|  | Overall | Care as Usual | Treatment |
|---|---|---|---|
| Overall | N=3125 | N=2,362 | N=763 |
| Sex | N (%) | N (%) | N (%) |
| Female | 2,035 (65%) | 1,539 (65%) | 496 (66%) |
| Male | 1,090 (35%) | 823 (35%) | 267 (38%) |
| Age group |  |  |  |
| 5-8 | 44 (1.4%) | 25 (1.1%) | 19 (2.5%) |
| 9-11 | 517 (17%) | 348 (15%) | 169 (22%) |
| 12-15 | 1,247 (40%) | 936 (40%) | 311 (41%) |
| 16-18 | 1,317 (42%) | 1,053 (45%) | 264 (35%) |
| Race/Ethnicity |  |  |  |
| Black | 1,713 (55%) | 1,341 (57%) | 372 (49%) |
| White | 807 (26%) | 599 (25%) | 208 (27%) |
| Hispanic/Latino | 426 (14%) | 288 (12%) | 138 (18%) |
| Another | 179 (5.7%) | 134 (5.7%) | 45 (5.9%) |
| Insurance at index |  |  |  |
| Medicaid | 1,747 (56%) | 1,375 (58%) | 372 (49%) |
| Private | 1,085 (35%) | 769 (33%) | 316 (41%) |
| Self-Pay | 230 (7.4%) | 168 (7.1%) | 62 (8.1%) |
| Other | 63 (2.0%) | 50 (2.2%) | 13 (1.7%) |
| Distance to Children’s |  |  |  |
| <50k | 2,717 (87%) | 2,036 (86%) | 681 (89%) |
| >50k | 408 (13%) | 326 (14%) | 82 (11%) |
| Preferred language |  |  |  |
| Spanish | 221 (7.1%) | 149 (6.3%) | 72 (9.4%) |
| English | 2,863 (92%) | 2,184 (92%) | 679 (89%) |
| Ano ther | 41 (1.3%) | 29 (1.2%) | 12 (1.6%) |
| Child Opportunity Index |  |  |  |
| Very Low | 890 (28%) | 702 (30%) | 188 (25%) |
| Low | 522 (17%) | 391 (17%) | 142 (19%) |
| Moderate | 570 (18%) | 456 (19%) | 114 (15%) |
| High | 482 (15%) | 357 (15%) | 125 (16%) |
| Very High | 650 (21%) | 456 (19%) | 194 (25%) |
| Emergency Department |  |  |  |
| Hughes Spaulding | 942 (30%) | 773 (33%) | 169 (22%) |
| Scottish Rite | 1,066 (34%) | 804 (34%) | 262 (34%) |
| Arthur Blank/Eggleston | 1,117 (36%) | 785 (33%) | 332 (44%) |

### Crisis Recovery Services Program versus Care as Usual

**Figure S1** shows the covariate balance plot for the intent-to-treat model before and after the application of inverse probability of treatment weights (IPTWs). Several demographic and clinical variables differed between treatment and care as usual groups prior to the application of the IPTWs, including insurance status, race/ethnicity, children opportunity index score, likelihood of the patient requiring a brief hospitalization, age group, and suicidal precaution level. Standard mean differences between the groups on all variables were reduced to less than 0.10 after the application of the weights (suggesting desirable covariate balance).

**Table 2** and **figure 1A** provide the regression analysis results for the 90-day intent-to-treat analysis. In this analysis, the treatment group included all children referred to crisis services regardless of whether they accessed those services. The mean number of ED revisits in 90 days in the treatment group was 0.10 (95%CI, 0.08-0.13) versus 0.17 (95%CI, 0.15-0.20) in the care as usual group. In the regression model, crisis services were associated with significantly reduced incidence of a mental health-related ED revisit within 90 days (adjusted incidence rate ratio (aIRR), 0.67; 95%CI, 0.56-0.81). The association between crisis services and reduced revisit rates was stronger in females (aIRR = 0.66; 95%CI, 0.52-0.84) than males (aIRR, 0.76; 95%CI, 0.57-1.01).

**Figure 1.**
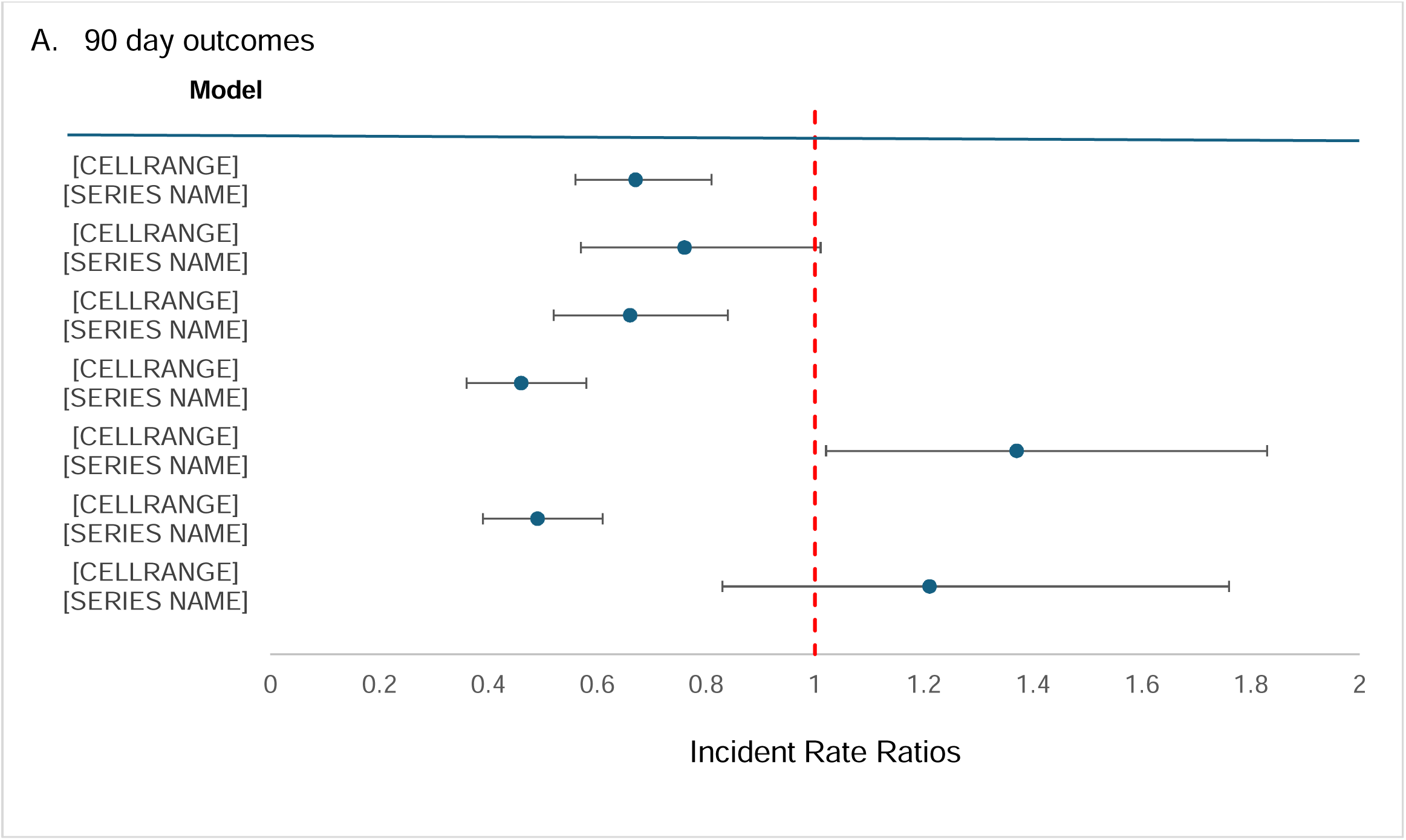
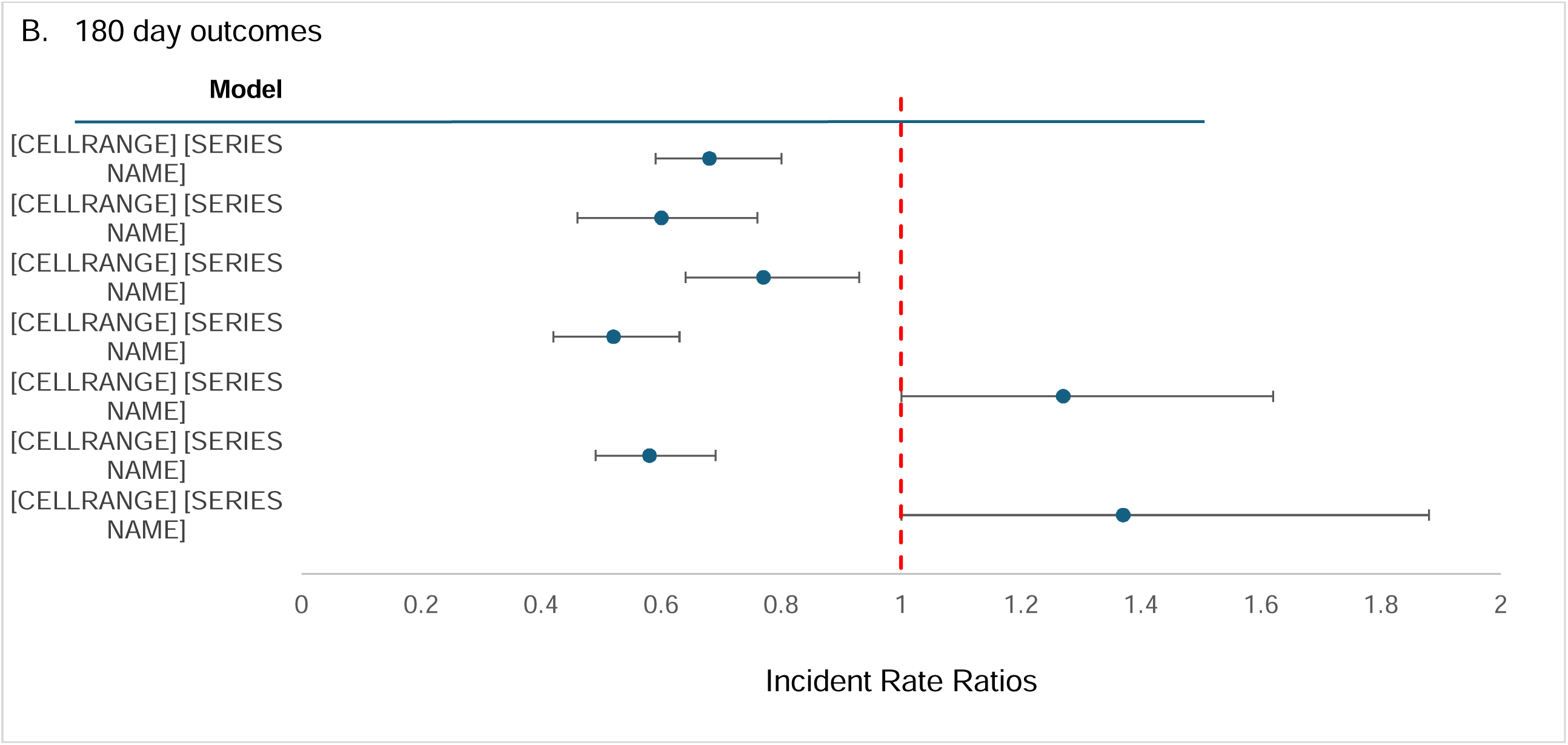
Forest plot of incidence rate ratios for intent-to-treat analysis at 90 days (A) and 180 days (B).

**Table 2.** Results of *intent-to-treat* analysis predicting the total number of ED revisits treated versus care as usual youth.

|  | 90 days |  |  |  |  |
|---|---|---|---|---|---|
|  | Total | Treatment | Care as Usual | Unadjusted | Propensity adjusted |
|  | N (Revisit #) | N (Revisit #) | N (Revisit #) | IRR 95%CI | IRR 95%CI |
| Overall ITT | 3,125 (485) | 763 (77) | 2,262 (408) | 0.58 (0.43, 0.79), <0.001 | 0.67 (0.56, 0.81), <0.001 |
| Males | 1,090 (161) | 267 (26) | 823 (135) | 0.59 (0.35, 0.99), 0.048 | 0.76 (0.57, 1.01), 0.056 |
| Females | 2,035 (222) | 496 (51) | 1,566 (273) | 0.58 (0.40, 0.84), 0.005 | 0.66 (0.52, 0.84), <0.001 |
| Hospitalized | 1,738 (341) | 157 (12) | 1,581 (329) | 0.37 (0.18, 0.70), 0.003 | 0.46 (0.36, 0.58), <0.001 |
| Not Hospitalized | 1,387 (144) | 606 (65) | 781 (79) | 1.06 (0.69, 1.62), 0.80 | 1.37 (1.02, 1.83), 0.05 |
| Red/Yellow level | 2,305 (400) | 430 (42) | 1,875 (358) | 0.51 (0.34, 0.75), <0.001 | 0.49 (0.39, 0.61), <0.001 |
| Green | 855 (97) | 345 (38) | 510 (59) | 1.21 (0.55, 1.65), 0.90 | 1.21 (0.83, 1.76), 0.3 |
|  | 180 days |  |  |  |  |
|  | N (Revisit #) | N (Revisit #) | N (Revisit #) | IRR 95%CI | IRR 95%CI |
| Overall ITT | 3,097 (743) | 759 (112) | 2,338 (631) | 0.55 (0.42, 0.71), <0.001 | 0.68 (0.59, 0.80), <0.001 |
| Males | 1,081 (250) | 265 (37) | 816 (213) | 0.53 (0.34, 0.82), 0.005 | 0.60 (0.46, 0.76), <0.001 |
| Females | 1,935 (493) | 494 (75) | 1,441 (418) | 0.55 (0.39, 0.77), <0.001 | 0.77 (0.64, 0.93), 0.007 |
| Hospitalized | 1,722 (522) | 157 (19) | 1,565 (503) | 0.38 (0.21, 0.66), <0.001 | 0.52 (0.42, 0.63), <0.001 |
| Not Hospitalized | 1,375 (221) | 602 (93) | 773 (128) | 0.93 (0.65, 1.33), 0.70 | 1.27 (1.00, 1.62), 0.052 |
| Red/Yellow level | 2,287 (622) | 430 (61) | 1,857 (561) | 0.47 (0.33, 0.66), <0.001 | 0.58 (0.49, 0.69), <0.001 |
| Green | 843 (139) | 340 (57) | 503 (82) | 1.03 (0.65, 1.62), 0.98 | 1.37 (1.00, 1.88), 0.052 |

All index visits were primarily for suicidal thoughts/behaviors. No MH ED visits in preceding 180 days; No preceding MH OP visits in the prior 180 days. Negative binomial regression was used to predict count of total ED revisits.

Treatment effects also varied based on acuity. First, a subset of qualifying ED visits resulted in transfer to inpatient psychiatric facility due to imminent risk (n = 1,738). Youth in this group were less likely to receive referrals to outpatient crisis services (only 9%), however, being in the treatment group significantly reduced the incidence of revisits within 90 days (aIRR, 0.46; 95% CI, 0.36-0.58). Conversely, in the group not transferred to a psychiatric facility, treatment was associated with a significantly *higher* incidence of ED revisits (aIRR, 1.37; 95% CI, 1.02-1.83). The second acuity-related variable was based on suicide precaution level. Youth at moderate or high suicide risk (i.e., yellow/red) had significantly reduced revisit incidence (aIRR, 0.49; 95% CI, 0.39-0.61), whereas youth with low suicide risk (i.e., green) did not display reduced ED revisit incidence (aIRR, 1.21; 95% CI, 0.83-1.76).

**Table 2** and **figure 1B** provide a similar series of results comparing the revisit incidence in both groups in the *180 days* following the index visit. In this longer timeframe, youth receiving crisis services continued to have a lower incidence of ED revisits (aIRR, 0.68; 95% CI, 0.59-0.80), and the effect was significant for both males and females. Reduced incidence of ED revisits was again only observed for youth with greater indicators of acuity (i.e., higher suicide precaution level and transfer to psychiatric facility).

### Sensitivity Analysis

Primary intent-to-treat models were retested with two subtypes of ED revisits: (1) revisits related to suicidality specifically and (2) revisits that resulted in transfer to inpatient psychiatric treatment. In both cases, referral to crisis services was associated with reduced incidence of these specific types of ED revisits at 90 days (suicide-related aIRR, 0.80; 95% CI, 0.64-1.01 and inpatient-related aIRR, 0.65; 95% CI, 0.52-0.81) and 180 days (suicide-related aIRR, 0.81; 95% CI, 0.67-0.98 and inpatient-related aIRR, 0.61; 95% CI, 0.51-0.74).

Next, all models were rerun under an as-treated framework which compared those that received at least 1 crisis service visit within 30 days to those that did not (e.g., care as usual group + referred only). As-treated analyses are useful for estimating the efficacy of the intervention apart from issues of non-adherence. **Table 3** and **figure 2** provide as-treated analysis results at 90 and 180 days. Results were similar or slightly stronger than those observed in the intent-to-treat analysis.

**Figure 2.**
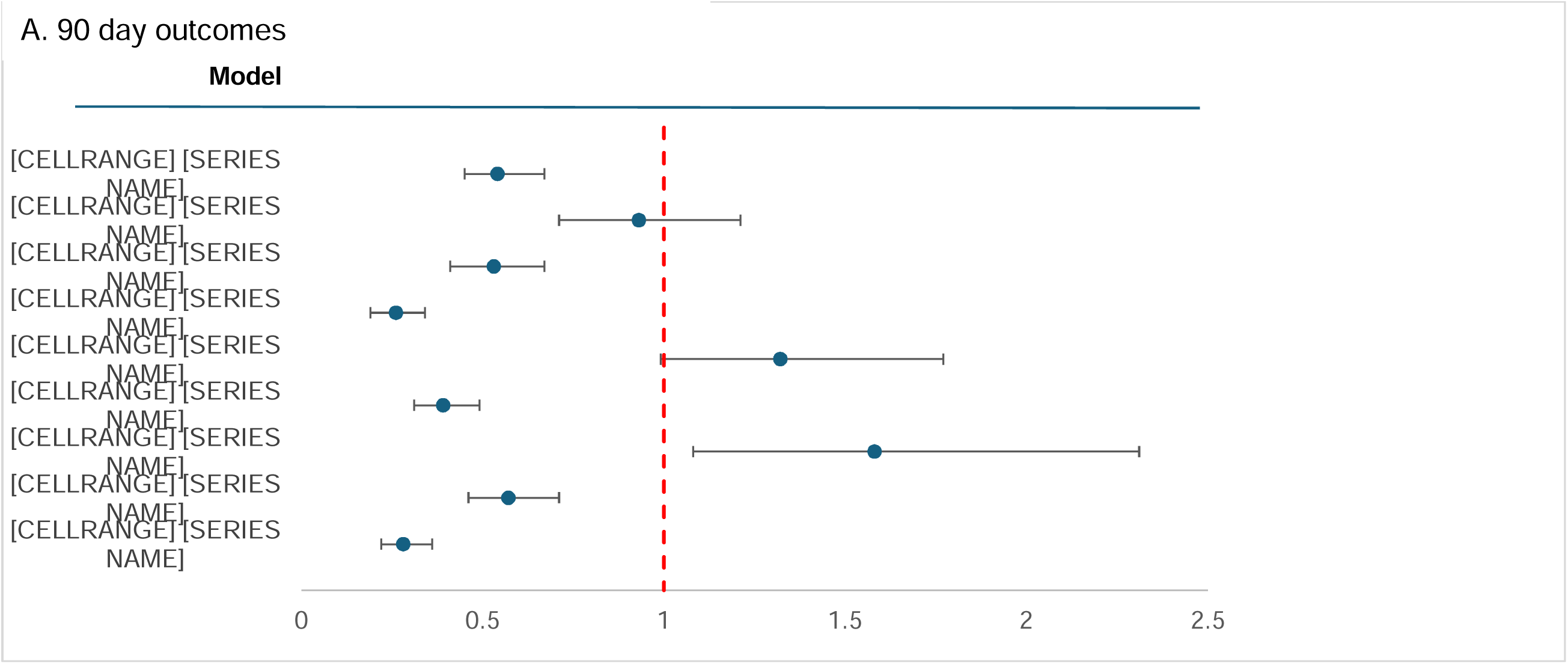
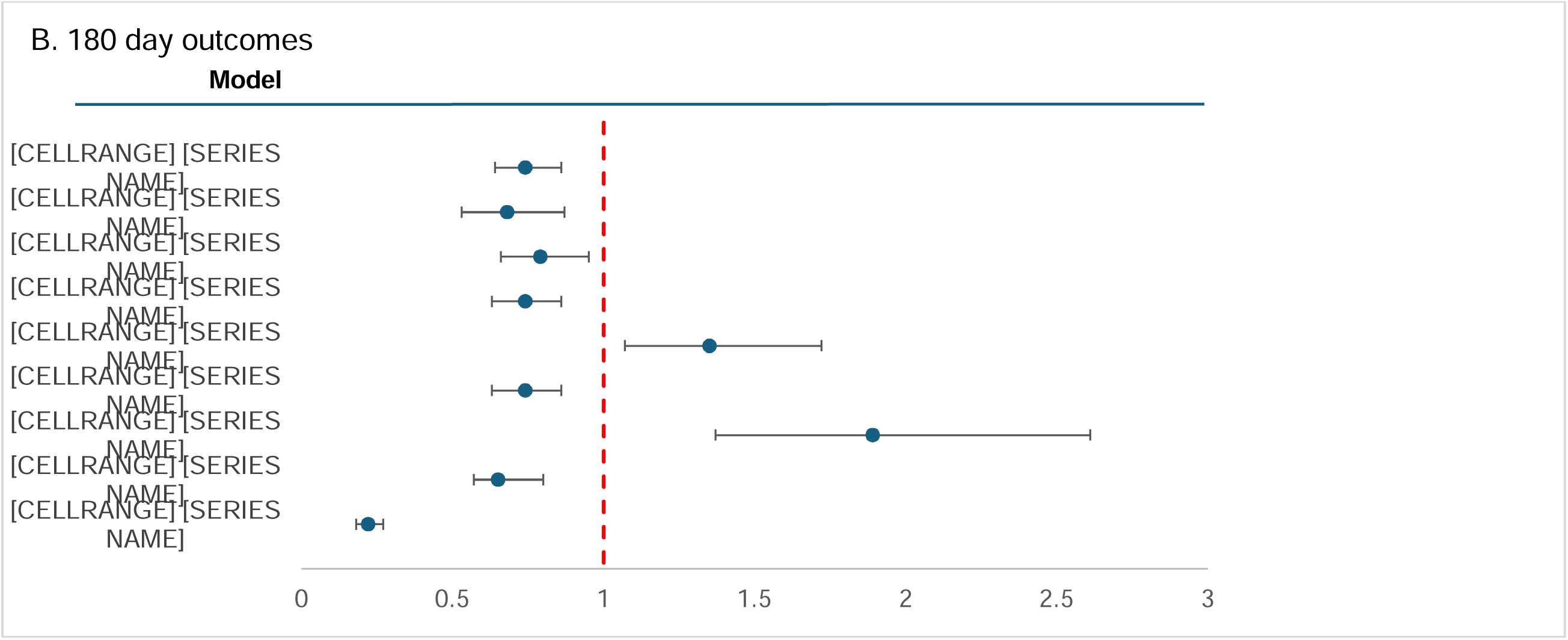
Forest plot of incidence rate ratios for as-treated analysis at 90 days (A) and 180 days (B).

**Table 3.** Results of *as-treated* analysis predicting the total number of ED revisits treated versus care as usual youth.

|  | 90 days |  |  |  |  |
|---|---|---|---|---|---|
|  | Total | Treatment | Care as Usual | Unadjusted | Propensity adjusted |
|  | N (Revisit #) | N (Revisit #) | N (Revisit #) | IRR 95%CI | IRR 95%CI |
|---|---|---|---|---|---|
| 1+ visit in 30 days | 3,125 (485) | 461 (50) | 2,664 (435) | 0.66 (0.46, 0.96), 0.030 | 0.55 (0.45, 0.67), <0.001 |
| Males | 1,090 (161) | 931 (17) | 781 (144) | 0.69 (0.36, 1.28), 0.20 | 0.93 (0.71, 1.21), 0.60 |
| Females | 2,035 (222) | 302 (33) | 1,733 (291) | 0.65 (0.41, 1.02), 0.065 | 0.53 (0.41, 0.67), <0.001 |
| Hospitalized | 1,738 (341) | 75 (6) | 1,663 (335) | 0.40 (0.14, 0.97), 0.054 | 0.26 (0.19, 0.34), <0.001 |
| Not Hospitalized | 1,387 (144) | 387 (44) | 1000 (100) | 1.14 (0.72, 1.81), 0.60 | 1.32 (0.99, 1.77), 0.058 |
| Red/Yellow level | 2,305 (400) | 252 (24) | 2,053 (376) | 0.52 (0.31, 0.85), 0.010 | 0.39 (0.31, 0.49), <0.001 |
| Green | 855 (97) | 189 (29) | 666 (68) | 1.27 (0.70, 2.32), 0.40 | 1.58 (1.08, 2.31), 0.020 |
| 2+ visits in 30 days | 2,962 (461) | 298 (26) | 2,664 (435) | 0.53 (0.33, 0.85), 0.010 | 0.57 (0.46, 0.71), <0.001 |
| 4+ visits in 30 days | 2,759 (444) | 95 (9) | 2,664 (435) | 0.58 (0.25, 1.26), 0.20 | 0.28 (0.22, 0.36), <0.001 |
|  | 180 days |  |  |  |  |
|  | N (Revisit #) | N (Revisit #) | N (Revisit #) | IRR 95%CI | IRR 95%CI |
| Overall ITT | 3,097 (743) | 461 (75) | 2,636 (668) | 0.46 (0.46, 0.88), 0.007 | 0.74 (0.64, 0.86), <0.001 |
| Males | 1,081 (250) | 159 (25) | 922 (225) | 0.64 (0.38, 1.08), 0.10 | 0.68 (0.53, 0.87), 0.002 |
| Females | 2,016 (493) | 302 (50) | 1,714 (443) | 0.64 (0.43, 0.96), 0.030 | 0.79 (0.66, 0.95), 0.011 |
| Hospitalized | 1,722 (522) | 75 (10) | 1,647 (512) | 0.43 (0.19, 0.92), 0.035 | 0.74 (0.63, 0.86), <0.001 |
| Not Hospitalized | 1,375 (221) | 386 (65) | 989 (156) | 1.17 (0.72, 1.57), 0.70 | 1.35 (1.07, 1.72), 0.013 |
| Red/Yellow level | 2,287 (622) | 252 (36) | 2,035 (586) | 0.50 (0.32, 0.76), 0.002 | 0.74 (0.63, 0.86), <0.001 |
| Green | 843 (139) | 340 (45) | 503 (94) | 1.40 (0.85, 2.29), 0.20 | 1.89 (1.37, 2.61), <0.001 |
| 2+ visits in 30 days | 2,934 (712) | 298 (44) | 2,636 (668) | 0.58 (0.39, 0.87), 0.008 | 0.65 (0.57, 0.80), <0.001 |
| 4+ visits in 30 days | 2731 (679) | 95 (11) | 2,636 (668) | 0.46 (0.21, 0.94), 0.039 | 0.22 (0.18, 0.27), <0.001 |

All index visits were primarily for suicidal thoughts/behaviors. No MH ED visits in preceding 180 days; No preceding MH OP visits in the prior 180 days. Negative binomial regression was used to predict count of total ED revisits.

## Discussion

This analysis of clinical program data couples implementation of an outpatient crisis clinic in a large health system with propensity models to test whether timely access to evidence-based crisis care could reduce ED recidivism for children experiencing suicidal thoughts and/or behaviors. Crisis services significantly reduced the incidence of ED revisits at both 3 months and 6 months post-index visit. Results varied based upon the acuity of the child’s symptoms and, to a lesser extent, by sex. Our results highlight the value of high-quality outpatient services for children in crisis and the importance of identifying which children are most likely to benefit and which may require alternative approaches.

Reductions in ED revisits were clinically significant, with a 33% and 32% reduction in the adjusted incidence rates of revisits at 3 months and 6 months, respectively. These findings contribute to the literature on effective approaches to reducing ED recidivism among children and adolescents with suicidality, an outcome for which prior systematic reviews have reported null effects in youth.^3,19,20^ This body of literature has examined ED-based interventions (e.g., implementing systematic follow-up contacts),^3,19,21^ family-based interventions,^19,20^ and community-based interventions (e.g., connecting to services in the community, equivalent to care as usual in our study).^22^ The significant and sizeable effect size of this current crisis program may be explained by several of its embedded features, including rapid access to treatment, outpatient space solely dedicated to crisis services, the availability of telehealth to increase service accessibility, clinicians trained in the delivery of crisis care, and the implementation of evidence-based interventions (including CAMS). Notably, the psychotherapy component of crisis services emphasizes dialectical behavior therapy (DBT) skills which have demonstrated moderate evidence for reducing suicidal ideation.^23^

The significant reduction in ED revisits was observed for those with the greatest acuity of symptoms (current suicidal ideation) and those requiring immediate transfer to an inpatient facility. This pattern has been observed with prior research on the CAMS interventions.^13^ Youth referred to crisis services with the least severe suicide risk experienced either no reduction in ED revisit incidence or – in some cases - an *increase* in ED revisits. There are several possible explanations for this result. First, crisis services primarily focused suicidality may have been a clinical mismatch for these children, especially if there were other pressing clinical concerns that went unaddressed.^24^ In addition, prior research has found that timely outpatient follow-up after a pediatric mental health ED visit may be associated with *increased* subsequent acute care use,^2^ as establishing a mental health provider may facilitate recognition of the need for referral to acute care services. Improving the match between risk assessment and admission to the crisis program may enhance the effectiveness and efficiency of available services.

Referral to crisis services reduced ED revisits among girls at 90 days (who account for most ED presentations for suicidality), but not among boys. This pattern may reflect differences in learning styles and level of engagement in services. Psychotherapeutic approaches like that implemented in CAMS may better fit the higher engagement levels in mental health interventions commonly observed among female versus male youth.^25,26^ A meta-analysis of 9 studies on suicidal adults participating in CAMS also reported that the effect sizes of the intervention were significantly reduced for male participants compared to the overall sample.^10^ These gender differences were not observed at 180 days, where both males and females showed benefit of intervention.

### Caveats

This evaluation tracked youths in one large urban health system, but findings are not necessarily representative of other large health systems or other geographic contexts. Similarly, this evaluation was completed in a health system in which clinicians were well-supported and maintained reasonable caseloads. Results may vary in other systems under different constraints.

Next, there were significant differences between our group in who was likely to receive a referral for crisis services before propensity score adjustment. Our conservative propensity-based approach approximates a randomized trial when all relevant covariates are measured and balanced, but there is still a potential for unmeasured confounding. Finally, while crisis services demonstrated beneficial effects on ED revisits – a significant outcome for any health system – the study did not evaluate other individual-, family-, or health system-level outcomes.

## Conclusion

This analysis highlights the promise of one health system’s effort to develop and implement a comprehensive, evidence-based approach to suicidality as a catastrophic aspect of the youth mental health crisis. The implementation of this program in a large urban hospital system was associated with a clinically significant reduction in ED recidivism among children who presented to the ED with suicidality. The effects of the approach differed by acuity of symptoms, with those experiencing the highest level of acuity particularly benefiting from the program. These findings highlight opportunities for learning health system approaches coordinated across national sites to continue to improve the effectiveness of outpatient recovery care in reducing the recurrence of suicidal crises of youth.

## Data Availability

The data of this study contain protected health information and are not publicly available. De-identified data may be made available from the corresponding author upon reasonable request and with appropriate institutional approvals.

## Abbreviations

ED: Emergency Department
LCSW: Licensed Clinical Social Worker
BMH: Behavioral Mental Health
aIRR: Adjusted Incidence Rate Ratio
MHS: Mental Health Specialist
CAMS: Collaborative Assessment and Management of Suicidality
ASQ: Ask Suicide-screening Questions

## What’s Known on This Subject

Emergency department visits for suicidal ideation and behavior among youth have increased substantially. Timely follow-up care is recommended, yet evidence remains limited regarding whether scalable outpatient crisis services can reduce subsequent mental health-related emergency department use.

## What This Study Adds

Referral to outpatient crisis services was associated with fewer mental health-related emergency department revisits at 90 and 180 days. Reductions were greatest among youth requiring short-term hospitalization and those with moderate-to-high suicide risk, suggesting particular benefit for higher-acuity youth.

## Contributors Statement Page

W. Copeland conceptualized and designed the study, carried out the analyses, drafted the initial manuscript, and critically reviewed and revised the manuscript.

J. Cumming, P. McCutcheon, H. Henderson J. Hudziak, and J. Constantino conceptualized and designed the study and critically reviewed and revised the manuscript.

N. Braykov harvested data from the EHR, designed the code for the primary analysis, and critically reviewed and revised the manuscript.

N. Muthu, X. Jing, A. Jahan, C. Ramos, L. Dilly, K. Krause, B. Livingston-Burns, L. Schneider, M. Graham, and G. Mukherjee critically reviewed and revised the manuscript.

All authors approved the final manuscript as submitted and agree to be accountable for all aspects of the work.

## Supplemental Materials

**Figure S1.**
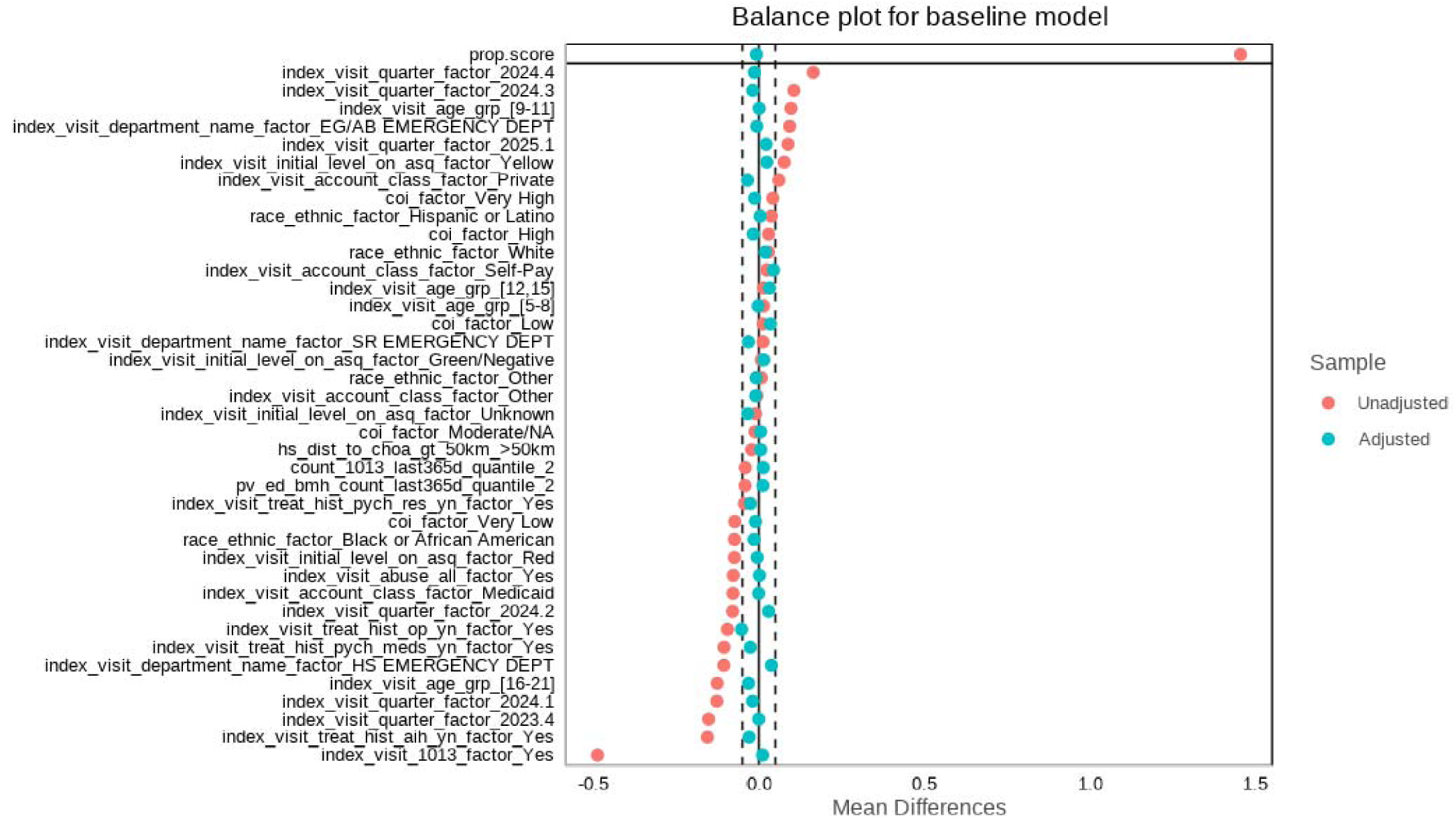

